# Management and Clinical Outcomes of Neonatal Hypothermia in the Newborn Nursery

**DOI:** 10.1101/2023.11.02.23297999

**Authors:** Rebecca Dang, Anisha I. Patel, Yingjie Weng, Alan R. Schroeder, Janelle Aby, Adam Frymoyer

**Author notes:** **Correspondence:** Rebecca Dang, Division of Pediatric Hospital Medicine, Lucile Packard Children’s Hospital Stanford, 453 Quarry Road, M/C 5776, Palo Alto, California 94304. **Role of Funder/Sponsor:** The Gerber Foundation had no role in the design and conduct of the study. Abbreviations: Lucile Packard Children’s Hospital Stanford (LPCHS); World Health Organization (WHO); Neonatal Intensive Care Unit (NICU); length of stay (LOS); electronic health record (EHR); small-for-gestational age (SGA); large-for-gestational age (LGA); infants of diabetic mothers (IDM); early-onset sepsis (EOS); C-Reactive Protein (CRP); Intravenous (IV); Newborn Weight Loss Tool (NEWT); gestational age (GA); birth weight (BW); Adjusted odds ratios (aOR); confidence intervals (CI).

## Abstract

**Background and Objectives:** Neonatal hypothermia has been shown to be commonly detected among late preterm and term infants. In preterm and very low birth weight infants, hypothermia is associated with increased morbidity and mortality. Little is known about the clinical interventions and outcomes in hypothermic late preterm and term infants. This study fills this gap in the evidence.

**Methods:** Single-center retrospective cohort study using electronic health record data on infants ≥35 weeks’ gestation admitted to a newborn nursery from 2015-2021. Hypothermia was categorized by severity: none, mild (single episode, 36.0-36.4°C), and moderate/severe (recurrent episodes and/or <36.0°C). Bivariable and multivariable logistic regression examined associations between hypothermia and interventions/outcomes. Stratified analyses by effect modifiers were conducted when appropriate.

**Results:** Among 24,009 infants, 1,111 had moderate/severe hypothermia. These hypothermic infants had higher odds of NICU transfer (aOR 2.10, 95% CI 1.68-2.60), sepsis evaluation (aOR 2.23, 95% CI 1.73-2.84), and antibiotic use (aOR 1.73, 95% CI 1.15-2.50) than infants without hypothermia. No infants with hypothermia had culture-positive sepsis and receipt of antibiotics ≥72 hours (surrogate for culture-negative sepsis and/or higher severity of illness) was not more common in hypothermic infants. Hypothermic infants also had higher odds of blood glucose measurement and hypoglycemia, higher percent weight loss and longer lengths of stay.

**Conclusion:** Late preterm and term infants with hypothermia in the nursery have potentially unnecessary increased resource utilization. Evidence-based and value-driven approaches to hypothermia in this population are needed.

**What’s Known on This Subject:** Neonatal hypothermia has been associated with morbidity and mortality in high-risk (preterm and very low birth weight) infants. The clinical implications of hypothermia in otherwise healthy late preterm and term infants admitted to the newborn nursery are poorly defined.

**What This Study Adds:** Infants with moderate/severe hypothermia have higher odds of diagnostic interventions and NICU transfers than infants without hypothermia. No infants with hypothermia had culture-positive sepsis. With the lack of a strong evidence base, hypothermia may drive unnecessary resource overutilization.

## INTRODUCTION

Neonatal hypothermia is common in the newborn nursery.^1–4^ In our prior study of >23,000 infants admitted to the nursery, we demonstrated that 21.7% infants had at least one episode of hypothermia,^4^ defined using the World Health Organization (WHO) threshold of <36.5°C.^5^ In high-risk preterm (≤33 weeks’ gestation) and/or very low birth weight (<1,500 g) infants admitted the Neonatal Intensive Care Unit (NICU), hypothermia has been associated with mortality and morbidity, including sepsis, intraventricular hemorrhage, bronchopulmonary dysplasia, necrotizing enterocolitis, and retinopathy of prematurity.^6–11^ However, little is known about the clinical outcomes of hypothermia in otherwise healthy late preterm and term infants admitted to the newborn nursery.

To our knowledge, the only published study evaluating clinical outcomes of hypothermia in the nursery is a retrospective chart review that demonstrated that hypothermia <36.5°C was associated with higher odds of NICU admission, need for respiratory support/diagnosis of respiratory distress syndrome, and increased length of stay (LOS).^1^ However, this study was limited by its small sample size (n=440 total infants; n=118 with hypothermia) and only evaluated hypothermia within 6 hours of birth. Given that late preterm and term infants represent over 3.5 million annual births in the US,^12^ it is essential to better understand healthcare resource utilization and outcomes for infants with hypothermia during their birth hospitalization in the nursery. The current study addresses this gap by evaluating diagnostic and therapeutic interventions and clinical outcomes associated with hypothermia in a much larger sample of late preterm and term infants admitted to a university-affiliated newborn nursery.

## METHODS

### Study Design

In this retrospective cohort study, we utilized electronic health record (EHR) data^13^ from infants and mothers admitted to the Lucile Packard Children’s Hospital Stanford (LPCHS) Newborn Nursery. The Stanford University Institutional Review Board approved this study.

### Setting

During the study period, the LPCHS Nursery had an annual birth volume of ∼4,500 infants and was located adjacent to a level II-IV NICU. Following delivery, healthy infants ≥35 weeks’ gestation and their mothers roomed-in together in the nursery. Due to a case of rectal perforation, routine rectal temperature measurement upon admission to the nursery was precautionarily discontinued before May 2015. Thereafter, only axillary temperatures (SureTemp Plus 692 or 6500 Connex Vital Signs Monitor Welch Allyn thermometers) were routinely measured. Rectal temperatures could be obtained per physician request. For temperatures <36.5°C, repeat axillary temperature and thermoregulatory measures (skin-to-skin contact, swaddling and/or additional clothing, and radiant warmer) were recommended. If temperature was still low following these interventions, a physician was notified. There were no formal hypothermia practice guidelines recommending specific management approaches. Other relevant clinical practices included routine glucose monitoring in small-for-gestational age (SGA), large-for-gestational age (LGA), and late preterm (35-36 ^6/7^ weeks) infants as well as infants of diabetic mothers (IDM), which aligns with the American Academy of Pediatrics Committee of Fetus and Newborn asymptomatic neonatal hypoglycemia practice guideline.^14,15^ LPCHS Nursery also employed an enhanced clinical observation approach to early-onset sepsis (EOS) screening through frequent clinical examinations.^16^

### Study Population

Our final cohort includes inborn infants admitted directly to the LPCHS nursery from the delivery room between May 2015 and August 2021. Patient flow is shown in Figure 1. Infants transferred to a non-NICU unit (i.e., acute care cardiac unit) or outside hospital were excluded, as were infants with no recorded temperature values or missing birth weight (due to a lack of comprehensive EHR documentation). We conducted data validation through random chart reviews on 110 infants in our final cohort and confirmed that all infants met inclusion criteria.

**Figure 1.**
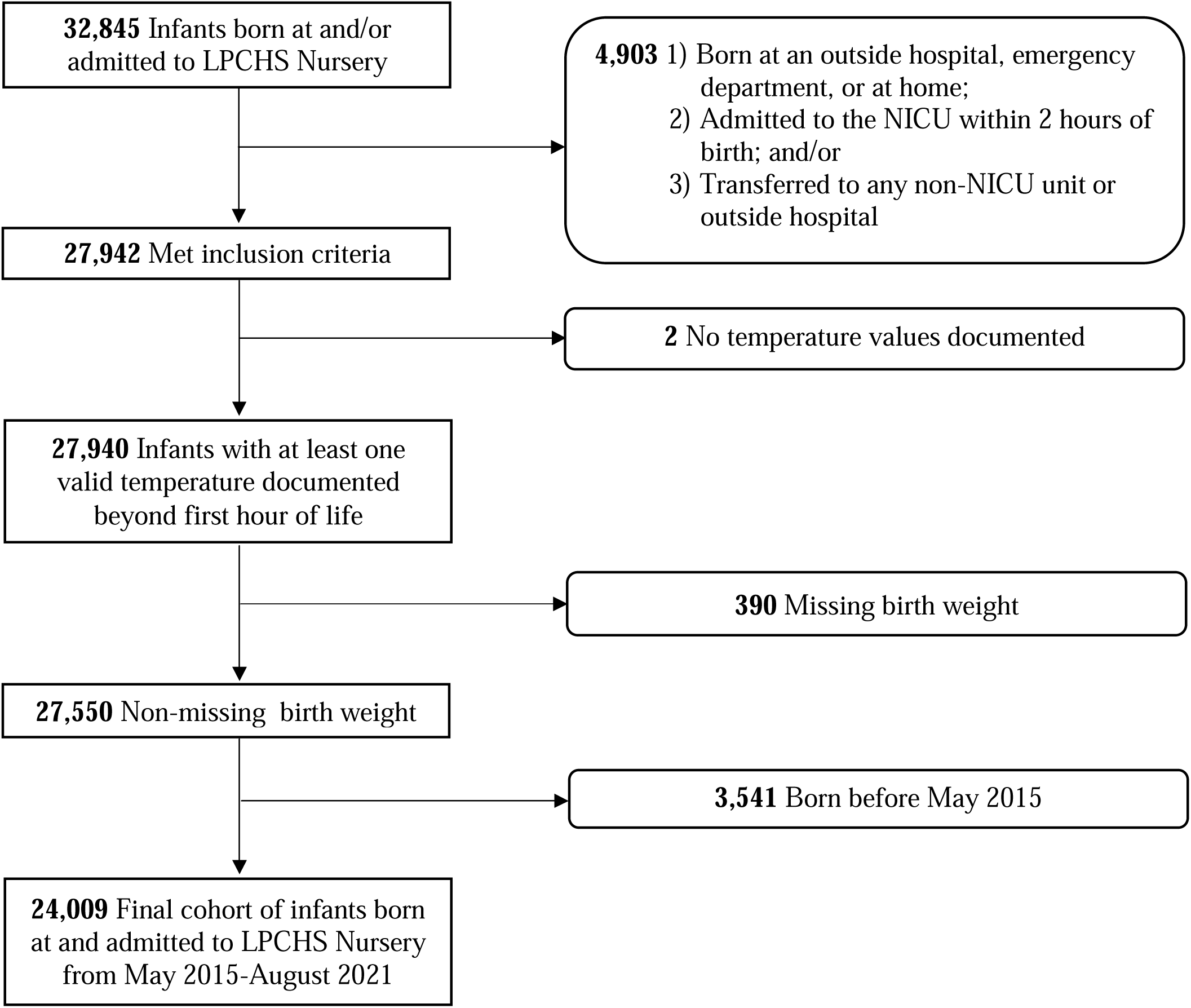
Patient Inclusion/Exclusion Flowchart From Data Extraction to Final Cohort.

### Outcomes

We *a priori* selected common interventions and clinical outcomes in infants with hypothermia for this study based on the research team’s clinical expertise and prior relevant literature.^1,5,9,11^ The primary outcome of study was NICU transfer, which was selected due to its important clinical and financial implications.^17–22^

#### Interventions

Secondary intervention-associated binary outcomes included 1) sepsis laboratory evaluations (C-Reactive Protein [CRP] and/or blood culture), 2) blood glucose measurement, 3) intravenous (IV) antibiotic receipt (ampicillin, gentamicin, cefotaxime, vancomyin, ceftazidime, and/or nafcillin), and 4) isolette therapy. If isolette bed type was documented concurrently with phototherapy, manual chart review was performed to determine whether isolette therapy was provided in response to hypothermia or just used to administer phototherapy. The latter scenario was not counted as isolette therapy.

#### Clinical Outcomes

Secondary clinically-associated outcomes: 1) blood culture-positive EOS, which did not include organisms commonly considered contaminants, including coagulase-negative *staphylococcus,*^23,24^ 2) prolonged antibiotic receipt ≥72 hours (as a proxy for culture-negative sepsis^25–28^ and/or “sicker” infant), 3) hypoglycemia (at least one blood glucose measurement of <35 mg/dL if <4 hours following birth or <45 mg/dL if ≥4 hours following birth), 4a) maximum percent weight loss during the period from birth until 72 or 96 hours after birth for vaginal and cesarean deliveries, respectively, or until discharge, whichever occurred first, 4b) proportion of infants with maximum percent weight loss above the vaginal or cesarean 75^th^ percentile curves on the Newborn Weight Loss Tool (NEWT),^29–31^ 5) length of stay (days), and 6) readmission to a Stanford-affiliated hospital within 14 days of nursery discharge.

### Exposures

Axillary and rectal temperatures after the first hour following birth were used to classify hypothermia status: no hypothermia (all temperature values ≥36.5°C), mild hypothermia (single episode lasting <2 hours, 36.0-36.4°C) and moderate/severe hypothermia (recurrent hypothermic episodes at least 2 hours apart or single episode <36.0°C). Thermometer routes documented as oral, tympanic, or temporal (i.e., routes not performed in the nursery) or missing (*n*=6,177, 1.9% of all analyzed values) were assumed to be inaccurately documented and were included in the analysis as axillary, the standard of care during the study period. Temperature outliers were identified by histogram and were all assessed through chart review. Temperature <34.0°C were deemed inaccurately documented in the EHR (e.g., no documentation of repeat temperature measurement, no mention in progress notes regarding the low temperature, and no interventions were pursued in response to the low temperature) and were therefore excluded (*n*=46).

### Statistical Analysis

Unadjusted rates of each intervention and clinical outcome by hypothermia category were compared in bivariable and multivariable analysis. Given known clinical differences between late preterm and term infants,^32–34^ gestational age (GA; defined by completed weeks, i.e., 35 weeks, 36 weeks, 37 weeks, etc.) was included as a confounder for all interventions and outcomes except for weight loss (adjusted for mode of delivery: cesarean or vaginal) and glucose measurement/hypoglycemia (adjusted for documented risk factors for hypoglycemia: SGA, LGA, and late preterm). Tachypnea (respiratory rate ≥60 breaths per minute documented ≥4 hours apart^35^), infant fever (≥38.0°C), and maternal fever (≥38.0°C during labor through 1 hour postpartum) were tested as effect modifiers of sepsis laboratory evaluation, IV antibiotic receipt, and transfer to the NICU. If an effect modifier was found to be statistically significant, a stratified analysis was conducted using generalized linear regression. As isolette therapy is provided for hypothermia, descriptive analyses were only performed in hypothermic infants.

Our prior manuscript on the epidemiology of hypothermia in the nursery found that both birth weight (BW) and GA were associated with hypothermia.^4^ Therefore, in the current study, we performed a sensitivity analysis adding BW as a confounder to all models that adjusted for GA. Missing data (0.05% for LOS and 2.7% for weight loss outcomes) were handled by complete case analysis. Adjusted odds ratios (aOR) with 95% confidence intervals (CI) were calculated. All analyses were conducted using R4.2.1 (R Foundation for Statistical Computing, Vienna, Austria).

## RESULTS

Of 32,845 infants born at LPCHS between May 2014 (the earliest date in the original database) and August 2021 (data extraction), 4,903 (14.9%) were excluded due to being 1) born at an outside hospital, in the emergency department, or at home; 2) admitted to the NICU within 2 hours of birth; and/or 3) transferred to an outside hospital or any unit other than the NICU (Figure 1). Of the remaining 27,942 included infants, 2 infants (0.01%) with no temperature value data and 390 (1.4%) with missing birth weight documentation were excluded. To better standardize temperature values and routes, 3,541 (12.9%) infants born before May 2015, when the temperature measurement protocol changed to discontinue routine rectal measurement, were excluded. The final cohort included 24,009 infants, of whom 4,080 (17.0%) had mild hypothermia and 1,111 (4.6%) had moderate/severe hypothermia. Moderate/severe hypothermia was comprised of 74.7% (830/1,111) with recurrent hypothermia, 14.2% (158/1,111) with a single episode of temperature <36.0°C, and 11.1% (123/1,111) with both. Demographic and clinical characteristics of infants by hypothermia status are shown in Table 1.

**Table 1.**
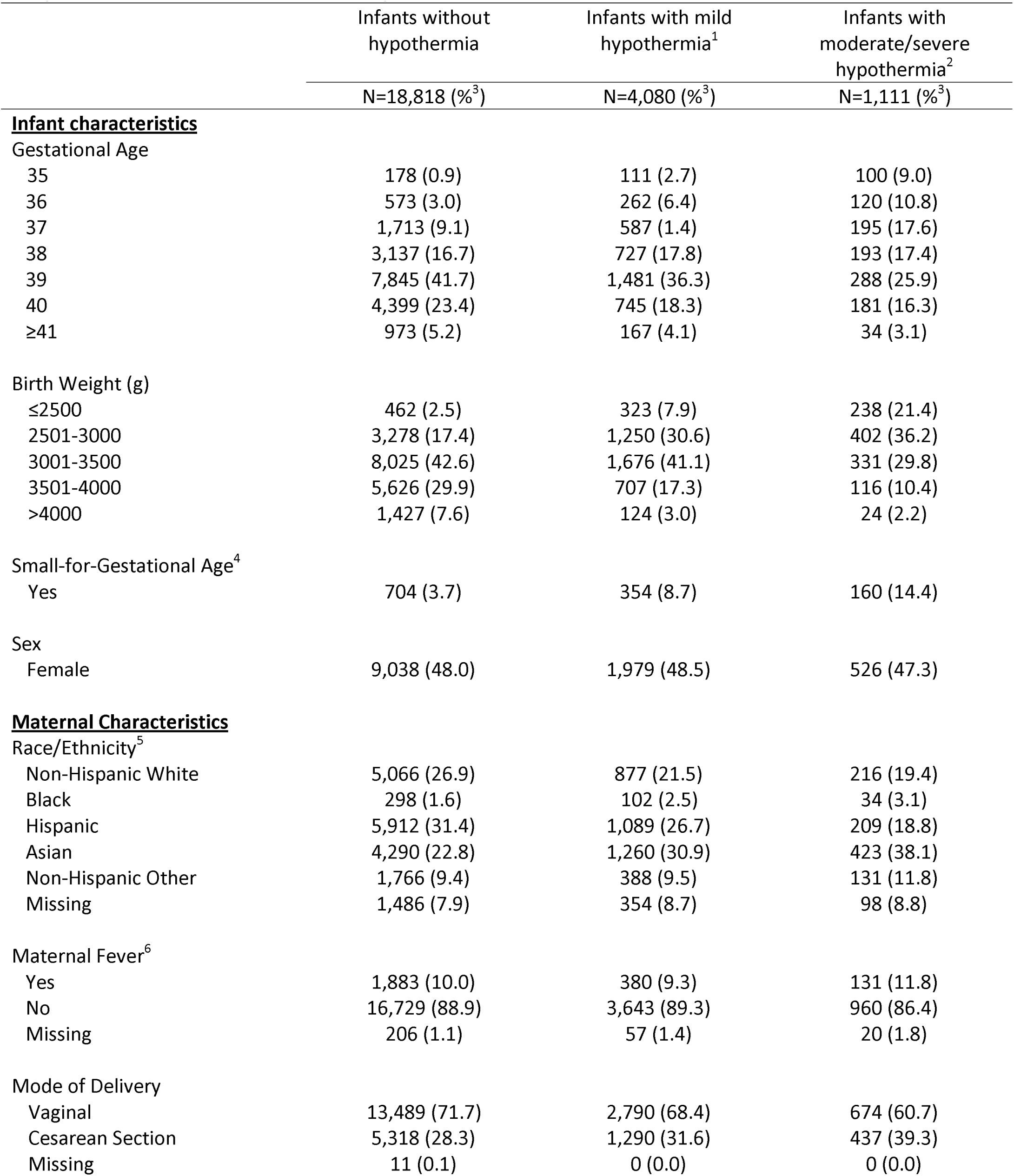
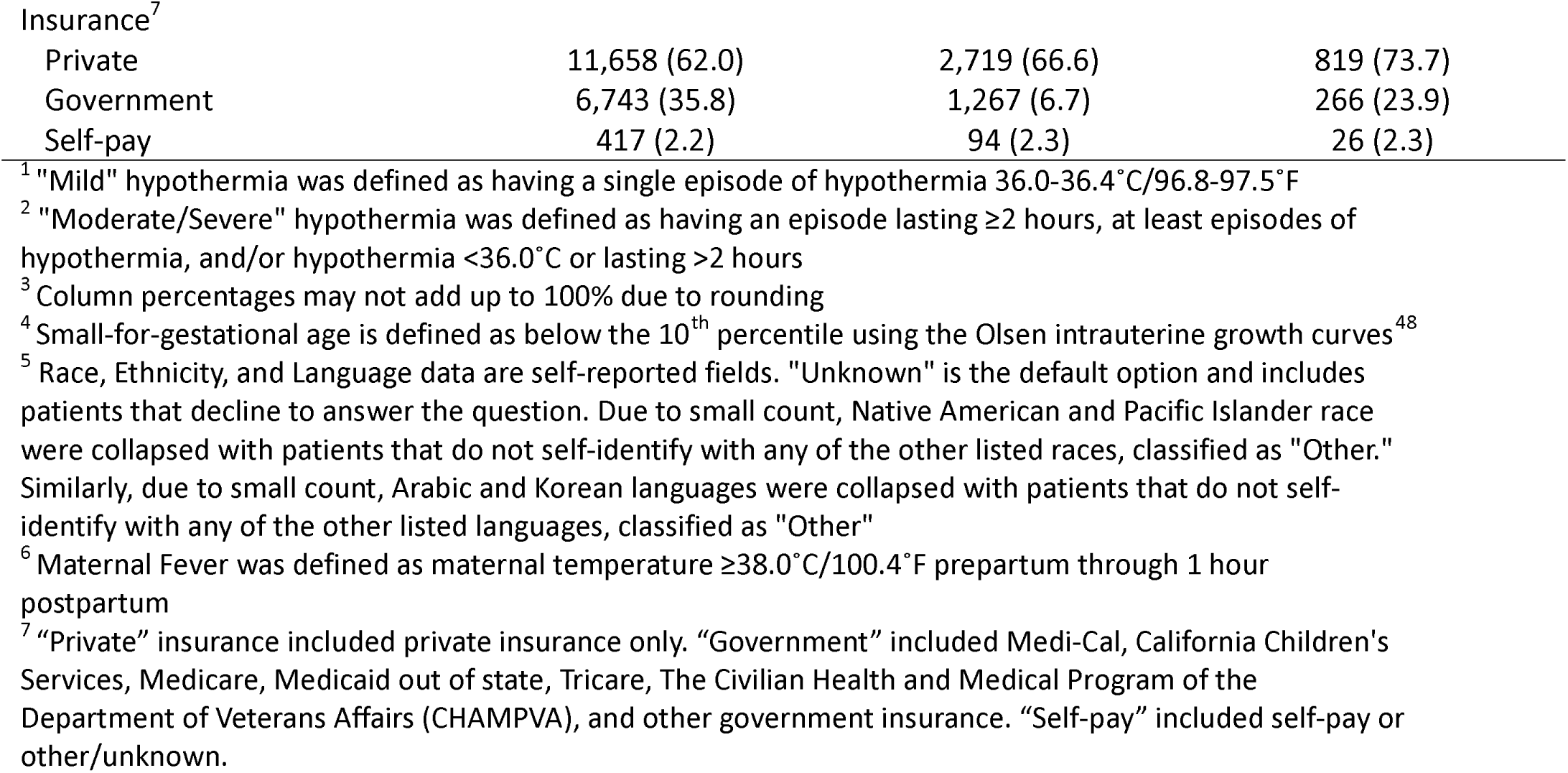
Characteristics of infants with and without hypothermia admitted to Lucile Packard Children’s Hospital Stanford (LPCHS) Newborn Nursery.

|  | Infants without hypothermia | Infants with mild hypothermia<sup>1</sup> | Infants with moderate/severe hypothermia<sup>2</sup> |
| --- | --- | --- | --- |
|  | N=18,818 (%<sup>3</sup>) | N=4,080 (%<sup>3</sup>) | N=1,111 (%<sup>3</sup>) |
| **<u>Infant characteristics</u>** |  |  |  |
| Gestational Age |  |  |  |
| 35 | 178 (0.9) | 111 (2.7) | 100 (9.0) |
| 36 | 573 (3.0) | 262 (6.4) | 120 (10.8) |
| 37 | 1,713 (9.1) | 587 (1.4) | 195 (17.6) |
| 38 | 3,137 (16.7) | 727 (17.8) | 193 (17.4) |
| 39 | 7,845 (41.7) | 1,481 (36.3) | 288 (25.9) |
| 40 | 4,399 (23.4) | 745 (18.3) | 181 (16.3) |
| ≥41 | 973 (5.2) | 167 (4.1) | 34 (3.1) |
| Birth Weight (g) |  |  |  |
| ≤2500 | 462 (2.5) | 323 (7.9) | 238 (21.4) |
| 2501-3000 | 3,278 (17.4) | 1,250 (30.6) | 402 (36.2) |
| 3001-3500 | 8,025 (42.6) | 1,676 (41.1) | 331 (29.8) |
| 3501-4000 | 5,626 (29.9) | 707 (17.3) | 116 (10.4) |
| >4000 | 1,427 (7.6) | 124 (3.0) | 24 (2.2) |
| Small-for-Gestational Age<sup>4</sup> |  |  |  |
| Yes | 704 (3.7) | 354 (8.7) | 160 (14.4) |
| Sex |  |  |  |
| Female | 9,038 (48.0) | 1,979 (48.5) | 526 (47.3) |
| **<u>Maternal Characteristics</u>** |  |  |  |
| Race/Ethnicity<sup>5</sup> |  |  |  |
| Non-Hispanic White | 5,066 (26.9) | 877 (21.5) | 216 (19.4) |
| Black | 298 (1.6) | 102 (2.5) | 34 (3.1) |
| Hispanic | 5,912 (31.4) | 1,089 (26.7) | 209 (18.8) |
| Asian | 4,290 (22.8) | 1,260 (30.9) | 423 (38.1) |
| Non-Hispanic Other | 1,766 (9.4) | 388 (9.5) | 131 (11.8) |
| Missing | 1,486 (7.9) | 354 (8.7) | 98 (8.8) |
| Maternal Fever<sup>6</sup> |  |  |  |
| Yes | 1,883 (10.0) | 380 (9.3) | 131 (11.8) |
| No | 16,729 (88.9) | 3,643 (89.3) | 960 (86.4) |
| Missing | 206 (1.1) | 57 (1.4) | 20 (1.8) |
| Mode of Delivery |  |  |  |
| Vaginal | 13,489 (71.7) | 2,790 (68.4) | 674 (60.7) |
| Cesarean Section | 5,318 (28.3) | 1,290 (31.6) | 437 (39.3) |
| Missing | 11 (0.1) | 0 (0.0) | 0 (0.0) |
| Insurance<sup>7</sup> |  |  |  |
| Private | 11,658 (62.0) | 2,719 (66.6) | 819 (73.7) |
| Government | 6,743 (35.8) | 1,267 (6.7) | 266 (23.9) |
| Self-pay | 417 (2.2) | 94 (2.3) | 26 (2.3) |

<sup>1</sup> "Mild" hypothermia was defined as having a single episode of hypothermia 36.0-36.4°C/96.8-97.5°F
<sup>2</sup> "Moderate/Severe" hypothermia was defined as having an episode lasting ≥2 hours, at least episodes of hypothermia, and/or hypothermia <36.0°C or lasting >2 hours
<sup>3</sup> Column percentages may not add up to 100% due to rounding
<sup>4</sup> Small-for-gestational age is defined as below the 10<sup>th</sup> percentile using the Olsen intrauterine growth curves<sup>48</sup>
<sup>5</sup> Race, Ethnicity, and Language data are self-reported fields. "Unknown" is the default option and includes patients that decline to answer the question. Due to small count, Native American and Pacific Islander race were collapsed with patients that do not self-identify with any of the other listed races, classified as "Other." Similarly, due to small count, Arabic and Korean languages were collapsed with patients that do not self-identify with any of the other listed languages, classified as "Other"
<sup>6</sup> Maternal Fever was defined as maternal temperature ≥38.0°C/100.4°F prepartum through 1 hour postpartum
<sup>7</sup> "Private" insurance included private insurance only. "Government" included Medi-Cal, California Children's Services, Medicare, Medicaid out of state, Tricare, The Civilian Health and Medical Program of the Department of Veterans Affairs (CHAMPVA), and other government insurance. "Self-pay" included self-pay or other/unknown.

### Interventions

NICU transfer, sepsis laboratory evaluation, and IV antibiotic receipt occurred in 4.2%, 3.1%, and 1.4% of all infants admitted to the nursery. Each intervention was more common in infants with moderate/severe hypothermia compared to infants without hypothermia (NICU transfer: aOR 2.10, 95% CI 1.68-2.60; sepsis laboratory evaluation: aOR 2.23, 95% CI 1.73-2.84; antibiotic receipt: aOR 1.73, 95% CI 1.15-2.50) (Table 2). When compared to infants without hypothermia, infants with mild hypothermia had statistically significant increased odds of NICU transfer but similar odds of sepsis laboratory evaluation and IV antibiotic receipt. When effect modifiers were tested, maternal fever was a statistically significant modifier of NICU transfer; tachypnea and infant fever were not. When stratified by presence of maternal fever, only hypothermic infants born to mothers without maternal fever had higher odds of NICU transfer. For sepsis laboratory evaluation and antibiotic receipt, tachypnea was a statistically significant modifier, but maternal and infant fever were not. In stratified analysis, higher odds of sepsis laboratory evaluation and antibiotic receipt in infants with moderate/severe hypothermia (and sepsis laboratory evaluation in infants with mild hypothermia) were only statistically significant in infants without tachypnea (Table 2).

**Table 2.** Bivariable and multivariable analyses of intervention-associated outcomes in infants admitted to the Lucile Packard Children’s Hospital Stanford (LPCHS) Newborn Nursery, by hypothermia severity.

|  | Infants without<br>hypothermia | Infants with mild<br>hypothermia<sup>1</sup> | Infants with<br>moderate/severe<br>hypothermia<sup>2</sup> |
| --- | --- | --- | --- |
|  | N=18,818 | N=4,080 | N=1,111 |
| **Primary Outcome** |  |  |  |
| **<u>NICU Transfer</u>** |  |  |  |
| Unadjusted Rates (%) | 657 (3.5) | 224 (5.5) | 126 (11) |
| Overall aOR (95% CI)<sup>3-4</sup> | Ref | **1.29 (1.10-1.51)** | **2.10 (1.68-2.60)** |
| Maternal fever aOR | Ref | 0.82 (0.46-1.37) | 1.29 (0.59-2.51) |
| No maternal fever aOR | Ref | **1.36 (1.14-1.61)** | **2.26 (1.79-2.84)** |
| **Secondary Outcomes** |  |  |  |
| **<u>Sepsis Laboratory Evaluation</u>** |  |  |  |
| Unadjusted Rates (%) | 515 (2.7) | 147 (3.6) | 88 (7.9) |
| Overall aOR (95% CI)<sup>3-4</sup> | Ref | 1.18 (0.97-1.42) | **2.23 (1.73-2.84)** |
| Tachypnea aOR | Ref | 0.89 (0.64-1.24) | 1.31 (0.82-2.10) |
| No tachypnea aOR | Ref | **1.55 (1.15-2.05)** | **3.38 (2.34-4.77)** |
| **<u>Antibiotic Receipt</u>** |  |  |  |
| Unadjusted Rates (%) | 236 (1.3) | 65 (1.6) | 33 (3.0) |
| Overall aOR (95% CI)<sup>3-4</sup> | Ref | 1.12 (0.84-1.47) | **1.73 (1.15-2.50)** |
| Tachypnea aOR | Ref | 1.16 (0.81-1.65) | 1.13 (0.67-1.88) |
| No tachypnea aOR | Ref | 0.89 (0.40-1.77) | **2.78 (1.22-5.70)** |
| **<u>Glucose Measurement (Y/N)</u>** |  |  |  |
| Unadjusted Rates (%) | 6,269 (33.3%) | 1,777 (43.6%) | 723 (65.1%) |
| aOR (95% CI)<sup>5</sup> | Ref | **1.43 (1.32-1.54)** | **2.94 (2.53-3.39)** |

<sup>1</sup> "Mild" hypothermia was defined as having a single episode of hypothermia 36.0-36.4°C/96.8-97.5°F
<sup>2</sup> "Moderate/Severe" hypothermia was defined as having an episode lasting ≥2 hours, at least episodes of hypothermia, and/or hypothermia <36.0°C or lasting >2 hours
<sup>3</sup> All adjusted analyses adjusted for gestational age
<sup>4</sup> Maternal fever, tachypnea, and infant fever were tested as effect modifiers for these outcomes. Only statistically significant modifiers are shown.
<sup>5</sup> Adjusted for infants with documented risk factors for hypoglycemia: small-for-gestational age, large-for-gestational age, and late preterm (35-36 weeks' gestation). While nursery protocol also included routine glucose measurement in infants of diabetic mothers, data on maternal diabetes were not available in this dataset.

Thirty-seven percent (8,769/24,009) of infants had at least 1 glucose measurement during their birth hospitalization. Compared to infants without hypothermia, infants with both mild (aOR 1.43, 95% CI 1.32-1.54) and moderate/severe (aOR 2.94, 95% CI 2.53-3.39) hypothermia had higher adjusted odds of glucose measurement (Table 2).

Less than 1 percent (0.4%; 18/4,080) of infants with mild hypothermia and 1.3% (14/1,111) with moderate/severe hypothermia were placed in an isolette. All hypothermic infants who received isolette therapy were ≤2,500 g and 71.9% (23/32) were late preterm. Twenty-five of the 32 infants who received isolette therapy also received IV antibiotics.

### Clinical Outcomes

Of the 351 blood cultures obtained, there were only 3 cases of culture-positive EOS (all *Streptococcus agalactiae),* none of whom had hypothermia. Three other positive blood cultures were considered contaminants (i.e., antibiotics discontinued after speciation of coagulase-negative *staphylococci* and notes documented the culture as a contaminant). Prolonged receipt of antibiotics ≥ 72 hours was not associated with hypothermia (Table 3).

**Table 3.**
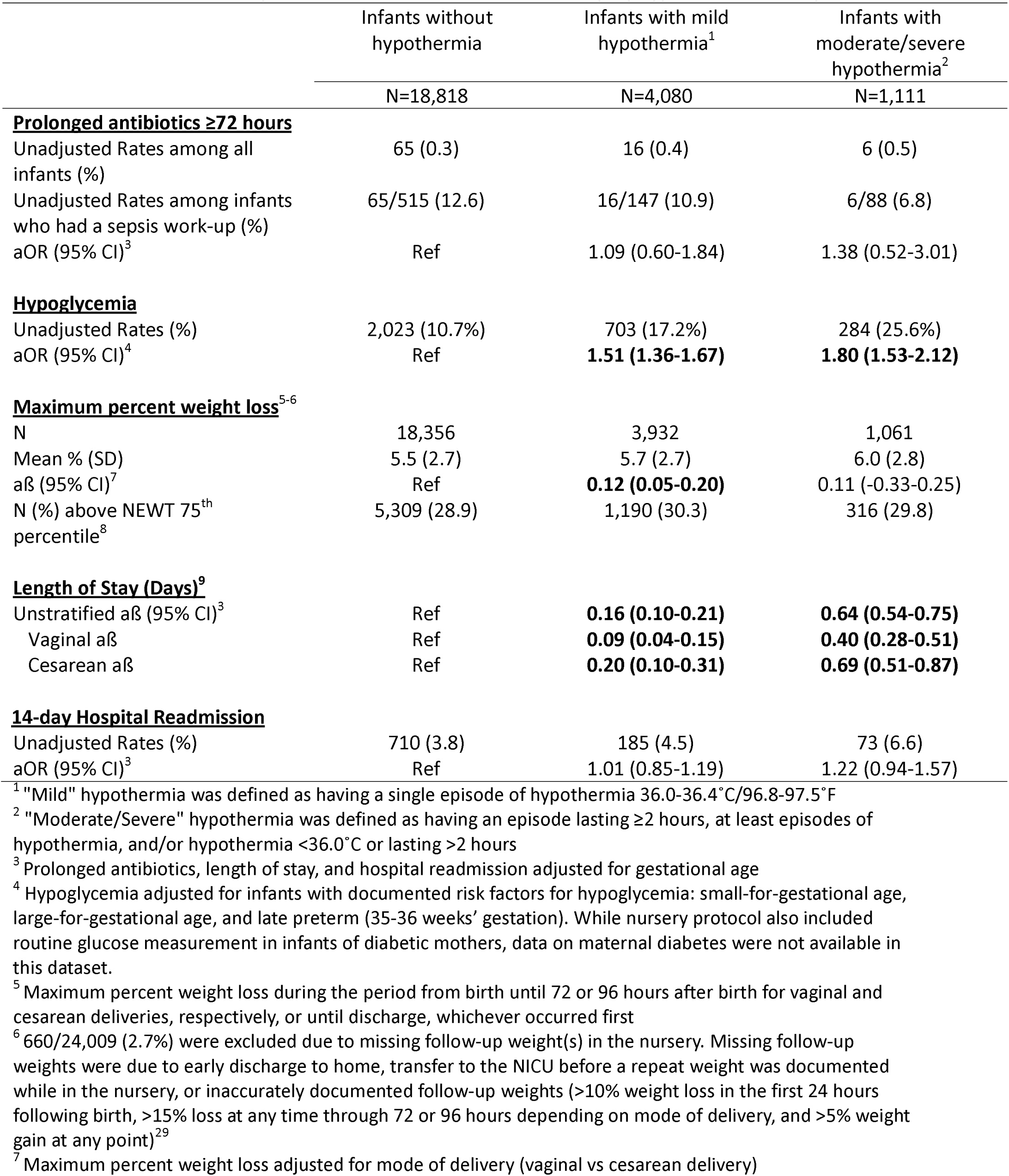
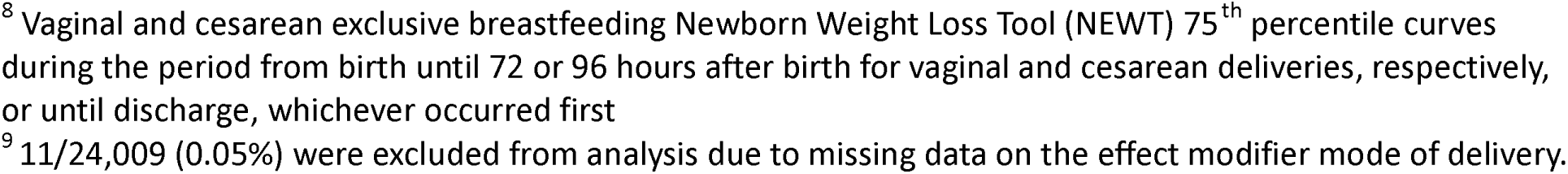
Bivariable and multivariable analyses of clinically-associated outcomes in infants admitted to the Lucile Packard Children’s Hospital Stanford (LPCHS) Newborn Nursery, by hypothermia severity.

Infants with hypothermia had higher adjusted odds of hypoglycemia than infants without hypothermia, which increased by hypothermia severity (mild: aOR 1.51, 95% CI 1.36-1.67; moderate/severe: aOR 1.80, 95% CI 1.53-2.12) (Table 3).

Hypothermia was associated with higher weight loss and longer LOS, but not hospital readmission (Table 3). The maximum percent weight loss adjusted for mode of delivery was 0.12 (95% CI 0.05-0.20)% higher in infants with mild hypothermia than without hypothermia; the higher percent weight loss was not statistically significant in infants with moderate/severe hypothermia. LOS was 0.16 (95% CI 0.10-0.21) and 0.64 (95% CI 0.54-0.75) days longer in infants with mild and moderate/severe hypothermia, respectively. These associations remained when stratified by mode of delivery (Table 3).

### Sensitivity Analysis

Adjusting for BW and GA yielded similar findings as the primary analysis (Supplement A).

## Discussion

This study of over 24,000 late preterm and term infants is the largest study to date investigating the clinical implications of neonatal hypothermia in the newborn nursery. We found that infants with hypothermia had higher odds of interventions including NICU transfer, sepsis laboratory evaluation, antibiotic receipt, and glucose measurement than infants without hypothermia.

However, there were no cases of culture-positive sepsis in over 5,000 infants with hypothermia, and hypothermia did not confer higher odds of prolonged antibiotic therapy (i.e., culture-negative sepsis and/or a “sicker” infant). Hypothermic infants did have higher odds of hypoglycemia and weight loss than non-hypothermic infants. Our data illustrate that the relatively common finding of hypothermia in the nursery may contribute to increased resource utilization. The harm-benefit profiles of interventions to assess for and treat hypothermia warrant further evaluation.

In the current study, infants with hypothermia had higher odds of NICU transfer (mild: aOR 1.29, 95% CI 1.10-1.51; moderate/severe: aOR 2.10, 95% CI 1.68-2.60) and longer LOS (mild: +0.16 days, 95% CI 0.10-0.21; moderate/severe: +0.64 days, 95% CI 0.54-0.75) than infants without hypothermia. When stratifying NICU transfer by maternal fever, the association with hypothermia was only demonstrated in infants without maternal fever. This suggests that hypothermia may increase the risk for NICU transfer in healthy infants but may not add significant additional risk in infants already at high risk for NICU transfer (i.e., infants born to mothers with peripartum fever). A smaller prior study (n=440; n=118 with hypothermia within 6 hours of birth) of infants ≥34 weeks’ gestation also demonstrated higher odds of NICU transfer (OR 2.87, 95% CI 1.36-6.04) and longer LOS (1.20 times longer, 95% CI 1.04-1.37) in hypothermic infants compared to non-hypothermic infants.^1^ As NICU admission and longer LOS can increase cost^19–22^ and parental distress,^17,18^ future research should evaluate the most cost-effective and value-based care setting for late preterm and term infants with hypothermia.

Among the 5,191 hypothermic infants in our cohort, 235 underwent sepsis evaluations and 98 received antibiotics, with increased odds in healthy, non-tachypneic infants. None (95% CI 0.0- 0.06%) of these infants had culture-positive sepsis. Additionally, hypothermia was not associated with a prolonged antibiotic course ≥72 hours, which we used as a surrogate for culture-negative sepsis^25–28^ and increased severity of illness. Overall rates of EOS are low in late preterm (0.76- 1.0 cases per 1,000 infants) and term (0.31-0.5 cases per 1,000 infants) infants,^23,25,36^ including infants with hypothermia. Sepsis evaluation and empiric antibiotics in otherwise healthy hypothermic infants may not be warranted and could contribute to unnecessary harm, including excess “pokes” to the infant, mother-infant separation (and related effects such as increased parental depressive and anxiety symptoms^37–40^), increased antibiotic-resistance,^26,41^ disruption of normal gut microbiota,^42^ and healthcare resource overutilization.

Concern for hypoglycemia in hypothermic infants may also increase resource utilization. While there have been case reports highlighting low body temperature as an important clinical cue for hypoglycemia in adults,^43^ there are limited data on the relationship between hypothermia and hypoglycemia in newborns. A retrospective chart review of infants ≥34 weeks’ gestation (n=440, n=118 with hypothermia) found no association between hypothermia within 6 hours of birth and hypoglycemia within 12 hours of birth.^1^ Adjusting for documented hypoglycemia risk factors preterm, SGA, and LGA, our current study found increased odds of hypoglycemia in hypothermic infants compared to non-hypothermic infants, which increased with hypothermia severity (aOR 1.51 and 1.80 for mild and moderate/severe, respectively). We also found that the odds of glucose measurement increased by hypothermia severity (aOR 1.43 and 2.94 for mild and moderate/severe, respectively). It is challenging to disentangle a true relationship between hypothermia and hypoglycemia from overdiagnosis of hypoglycemia due to increased glucose measurement. A recent study showed 39% of otherwise healthy term infants who had routine repeat plasma glucose measurement performed experienced ≥ 1 episode of hypoglycemia (<47 mg/dL) in the first days of life,^44^ which supports that increased glucose measurement leads to increased hypoglycemia that would not otherwise have been detected. While hypoglycemia has known neurodevelopment impairments,^45,46^ unnecessary glucose measurement is not without harms and may negatively impact breastfeeding^47^ and contribute to increased asymptomatic hypoglycemia detection.^44^ Future multi-institution prospective research should further elucidate the added risk for hypoglycemia in infants with hypothermia to inform practice guidelines on the utility of glucose measurement when hypothermia is detected.

This study had limitations. First, we analyzed retrospective EHR data, which may not accurately capture all clinical events that occurred. To address this concern, we performed random chart review throughout the data extraction process to optimize data quality. Second, the retrospective study design lacks temporality and we were unable to differentiate whether hypothermia was driving interventions or if it was associated with other conditions that were driving interventions. To better understand the relationship between hypothermia and interventions/outcomes, we conducted a sensitivity analysis adding BW as a confounder and performed effect modification by clinical factors that may influence clinical decision making and intervention utilization. Third, while interventions and outcomes may have taken place prior to the first hypothermic event (≤1.0% in all analyses other than the glucose measurement and hypoglycemia analyses that included routine glucose screening), our findings demonstrate that infants with hypothermia during their birth hospitalization have higher healthcare resource utilization than infants without hypothermia. Finally, our study lacks generalizability as findings represent the unique population and specific practices encountered at LPCHS Nursery.

## Conclusions

Infants with moderate/severe hypothermia are at increased odds of NICU transfer, sepsis laboratory evaluation, antibiotic receipt, glucose monitoring and hypoglycemia, and longer length of stay compared to infants without hypothermia. However, these infants do not have evidence of a higher risk of EOS or need for prolonged antibiotic treatment. This study highlights that hypothermia contributes to high resource utilization in the nursery. Further investigation on the harm-benefit profiles of these approaches to the management, including preventive thermoregulatory practices, of hypothermia is necessary to inform high-value, evidence-based care surrounding neonatal hypothermia in the nursery.

## Data Availability

All data produced in the present study are available upon reasonable request to the authors

---

**Supplement A.** Sensitivity analysis examining impact of adding birthweight as a confounder in the multivariable analyses of clinically-associated outcomes in infants admitted to the Lucile Packard Children’s Hospital Stanford (LPCHS) Newborn Nursery, by hypothermia severity.

|  | Infants without<br>hypothermic<br>episodes<br>N=18,818 | Infants with mild<br>hypothermia<sup>1</sup> (95% CI)<br>N=4,080 | Infants with<br>moderate/severe<br>hypothermia<sup>2</sup> (95% CI)<br>N=1,111 |
| --- | --- | --- | --- |
| NICU Transfer aOR | Ref | **1.29 (1.09-1.52)** | **1.84 (1.45-2.30)** |
| Sepsis Laboratory Evaluation aOR | Ref | 1.21 (0.99-1.46) | **2.12 (1.63-2.74)** |
| Antibiotic Receipt aOR | Ref | 1.21 (0.90-1.60) | **1.84 (1.22-2.72)** |
| Prolonged antibiotics ≥72 hours aOR | Ref | 1.20 (0.66-2.06) | 1.48 (0.55-3.33) |
| Length of Stay (Days) aß | Ref | **0.15 (0.09-0.21)** | **0.56 (0.46-0.67)** |
| 14-day Hospital Readmission aOR | Ref | 1.03 (0.86-1.22) | 1.28 (0.98-1.66) |

<sup>1</sup> Birthweight was added as confounder for all analyses that adjusted for gestational age. All models presented adjust for birthweight and GA. Stratified analyses were not conducted due to insufficient power.
<sup>2</sup> "Mild" hypothermia was defined as having a single episode of hypothermia 36.0-36.4°C/96.8-97.5°F
<sup>3</sup> "Moderate/Severe" hypothermia was defined as having an episode lasting ≥2 hours, at least episodes of hypothermia, and/or hypothermia <36.0°C or lasting >2 hours

